# Unearthed Novel Molecular Phenotypes and Potential Therapeutic Targets in Esophagogastric Adenocarcinoma

**DOI:** 10.1101/2024.07.08.24310077

**Authors:** Annika Windon, Majd Al Assaad, Kevin Hadi, Nicole Mendelson, Erika Hissong, Aditya Deshpande, Marvel Tranquille, Justin Mclee, Minal Patel, Juan S. Medina-Martínez, Kenrry Chiu, Jyothi Manohar, Michael Sigouros, Allyson J. Ocean, Andrea Sboner, José Jessurun, Olivier Elemento, Manish Shah, Juan Miguel Mosquera

**Affiliations:** Department of Pathology and Laboratory Medicine, Weill Cornell Medicine, New York, NY, USA; Englander Institute for Precision Medicine, Weill Cornell Medicine, New York, NY, USA; Isabl Inc., New York, NY, USA; Department of Medicine, Division of Hematology and Medical Oncology, Weill Cornell Medicine, New York, NY, USA; Institute for Computational Biomedicine, Weill Cornell Medicine, New York, NY, USA

**Keywords:** Esophageal adenocarcinoma, gastric adenocarcinoma, whole genome sequencing, molecular targets, oncology biomarkers

## Abstract

**Background:** Esophagogastric adenocarcinoma demands a deeper molecular understanding to advance treatment strategies and improve patient outcomes. Here, we profiled the genome and transcriptome landscape of these cancers, explored molecular characteristics that are invisible to other sequencing platforms, and analyzed their potential clinical ramification.

**Methods:** Our study employed state-of-the-art analyses of whole genome and transcriptome sequencing on 52 matched tumor and germline samples from 47 patients, aiming to unravel new therapeutic targets and deepen our understanding of these cancers’ molecular foundations.

**Results:** The analyses revealed 88 targetable oncogenic mutations and fusions in 62% of the patients, and further elucidated molecular signatures associated with mismatch repair and homologous recombination deficiency. Notably, we identified *CDK12*-type genomic instability associated with *CDK12* fusions, novel *NTRK, NRG1, ALK,* and *MET* fusions, and structural variants in relevant cancer genes like *RAD51B*.

**Conclusions:** Our findings demonstrate the power of integrative whole genome and transcriptome sequencing in identifying additional therapeutic targets, supporting a promising path for precision medicine in treating esophagogastric adenocarcinoma.

## Background

Malignancies of the gastroesophageal junction (GEJ) and stomach remain prevalent in the United States and worldwide. The American Cancer Society predicts more than 26,000 new cases and 10,000 deaths in the United States in 2024 (1). According to the Surveillance, Epidemiology, End Results (SEER) database, the 5-year survival rate for gastric cancer is 36%, emphasizing its dismal prognosis(2). Risk factors for gastric adenocarcinoma include *Helicobacter pylori* infection, tobacco smoking, high salt intake, and family history (3). Proximal (cardiac-type) and GEJ adenocarcinoma share different risk factors including Barrett esophagus, male sex, age over 50 years, gastroesophageal reflux disease, and obesity (3–5).

Genomic research in esophagogastric adenocarcinomas (EGAC) has revealed predominant driver mutations in genes such as *TP53, CDKN2A, SMAD4, ARID1,* and *PIK3CA* with targetable therapy available for genetic alterations in pathways involving *MET, PIK3CA, EGFR, ERBB2, and ERBB3* (6–9). Presently, there are three main biomarkers that predict improved treatment response to targeted therapy for EGAC which include microsatellite instability (MSI) status, HER2 positivity, and PD-L1 expression (6). Apart from the progress in HER-2 targeted therapy for EGAC (10, 11), advancements in conventional chemotherapy, both as an adjunct to surgery and in a curative context, have been minimal(12). Recent publications and ongoing clinical trials targeting PD-L1 and adenocarcinoma with high microsatellite instability (MSI-H), including checkmate 577, NEONIPIGA, VESTIGE, and KEYNOTE-585, indicate that immunotherapy is yielding encouraging results, either as a standalone treatment or in conjunction with chemotherapy(12–16). Despite these known biomarkers, there is still an unmet clinical need to find better molecular targets and further guide the development of novel therapeutic agents for EGAC. In our study, we conducted an integrative analysis of whole genome and transcriptome sequencing (WGTS) on a cohort of EGAC patients to unearth novel biomarkers and explore their potential impact on targeted therapy.

## Methods

### Patient Samples and Pathologic Examination

Patients were enrolled at Weill Cornell Medicine (WCM) under an institutional review board (IRB)– approved protocol (Research for Precision Medicine WCM IRB No. 1305013903) with written informed consent. Retrospective tissue samples (year of collection 2014 - 2021) were retrieved and studied under the protocol for Comprehensive Cancer Characterization by Genomic and Transcriptomic Profiling (WCM IRB No. 1007011157). Patient demographics, primary site, tumor site, pathological grade, and treatment history were obtained from the pathology reports and electronic medical records (EMR). Samples in this study included adenocarcinoma from the following primary sites: distal esophagus, GEJ, and stomach and are collectively referred to as esophagogastric adenocarcinoma (EGAC). Representative hematoxylin and eosin-stained slides from each case were reviewed by the study pathologists.

### Whole Genome Sequencing, RNA-Sequencing and Integrative Data Analysis

Whole-genome sequencing (WGS) was conducted on paired tumor and normal samples as described previously(17–19). In summary, DNA was extracted from both tumor and normal tissues and the DNA quality and amount were evaluated using the Agilent Tapestation 4200 (Agilent Technologies) and Qubit Fluorometer (ThermoFisher). WGS libraries were constructed with the KAPA Hyper Library Preparation Kit (KAPABiosystems KK8502, KK8504), and the DNA fragments were then end-repaired, adenylated, connected to Illumina sequencing adapters, size-selected using beads, and amplified. Sequencing was carried out at the New York Genome Center on an Illumina Novaseq6000 sequencer with 2×150bp cycles. Sequencing data were mapped to the hg19 reference genome using the Burrows-Wheeler aligner (20). The entire alignment and variant identification process adhered to GATK best practices. Sample contamination and concordance were checked using Conpair(21). The Isabl GxT analytical platform was utilized(22, 23). We curated single nucleotide variants (SNVs), insertions/deletions (Indels), and structural variants (SVs) linked to tumor suppressor genes and oncogenes(17, 22, 24, 25). In addition to single base substitution (SBS) molecular signatures, whole genome and coding region tumor mutational burden (TMB), microsatellite instability (MSI), and homologous recombination deficiency (HRD) were also curated. TMB ≥10 mutations/megabase (mut/mb) was considered TMB-high (TMB-H) and MSI ≥10 was considered MSI-high (MSI-H).

RNA sequencing (RNA-seq) and its corresponding data analysis were carried out as detailed in prior studies (26, 27). In brief, total RNA was isolated followed by cDNA library preparation. Pair-end RNA-seq was performed on GAII, HiSeq 2000, or HiSeq 2500 platforms(26, 27). The sequencing reads were independently aligned against the human genome sequence build hg19 using STAR_2.4.0f1(21). The expression levels (FPKMS) were estimated using Cufflinks (2.0.2), and the GENCODE v19 GTF file was employed for annotation(28). Integration of RNA-seq results with WGS data was visualized in an interactive web-based interface portal (19). The Isabl GxT analytic platform is linked to the Memorial Sloan Kettering Cancer Center’s Precision Oncology Knowledge Base (OncoKB)(29), which provides relevance of detected variants.

## Results

### Clinical and Pathologic Characteristics of the Study Cohort

The cohort comprised 52 samples of EGAC from 47 patients with a median age of 65.9 years (range: 24.7-88.1 years) and a male predominance (**Table 1**). Twenty-three samples were obtained from primary tumor sites, and 29 samples were from metastatic sites including the peritoneum, liver, lymph nodes, bone, and brain. Fourteen tumor DNA samples were obtained from formalin-fixed, paraffin-embedded (FFPE) tissue, 36 from frozen tissue, and 2 from ascitic fluid. Germline DNA from all 47 patients was extracted from blood samples. RNA, available for 28 samples was extracted from the same tissue sections as the DNA. Twenty-two samples had a distal esophageal/gastroesophageal junction origin, and 30 samples were classified as gastric in origin.

### Integrative Analysis of WGS and RNAseq Elucidates Molecular Targets and Relevant Biomarkers in EGAC including Structural Variants Impacting Cancer Genes

A total of 2,633 alterations in cancer-related (oncogenes or tumor suppressor) genes were identified. These alterations comprised 146 germline variants, 672 copy number variants (CNVs), 720 small mutations (including Indels), and 1,095 structural variants (SVs). A summary of molecular alterations is illustrated in **Figure 1**.

**Figure 1.**
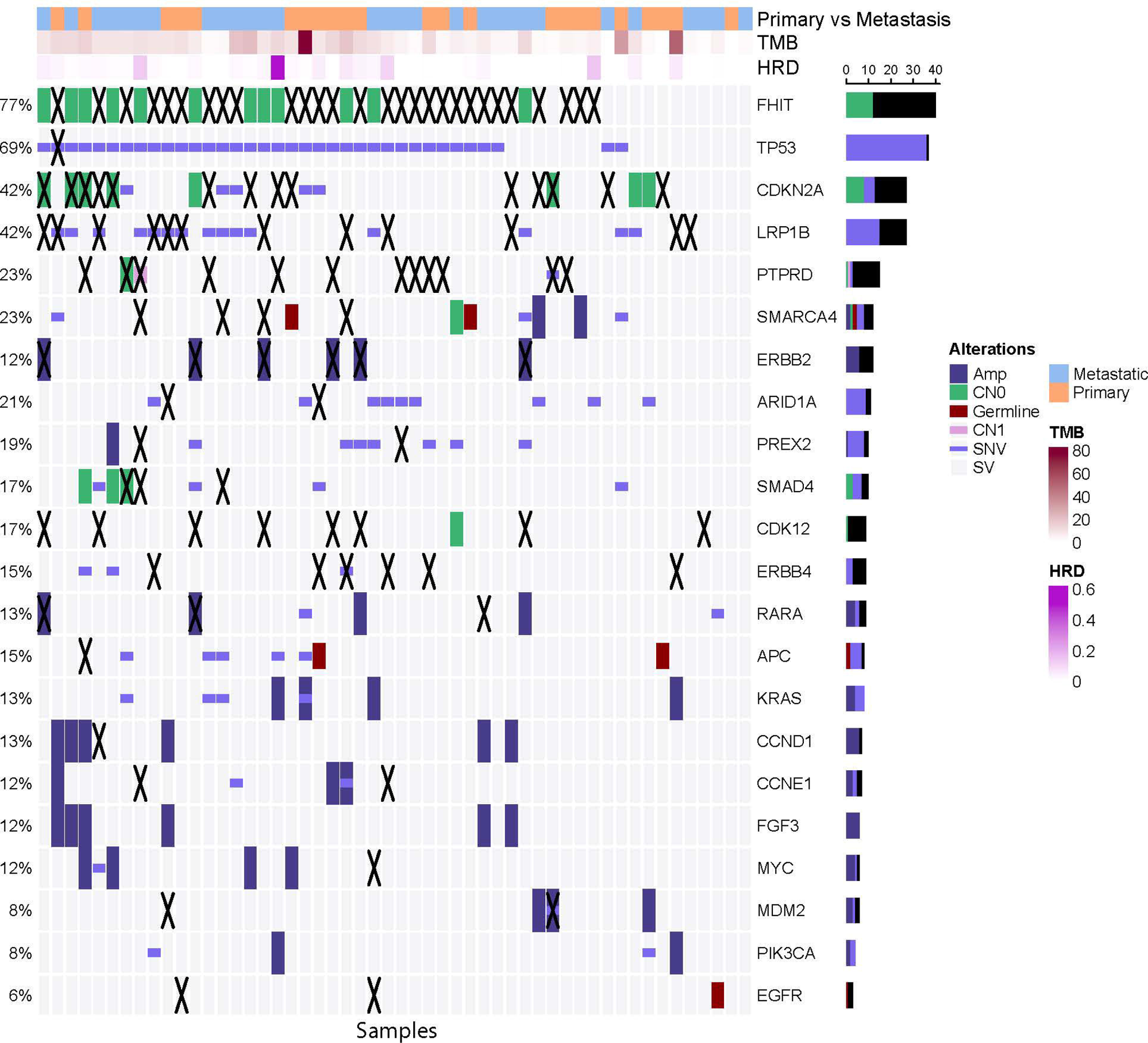
Whole genome and transcriptome landscape of esophagogastric adenocarcinoma. Oncoplot illustrates the spectrum of somatic, germline, and structural variants in cancer genes, as well as HRD and TMB biomarkers. Abbreviations: HRD = Homologous recombination deficiency; TMB = Tumor mutational burden; Amp = Amplification; CN1 = Copy number 1; SV = Structural variant; SNV = Single nucleotide variant.

We explored the presence of SVs impacting genes known to be drivers of EGAC. Such SVs included complex rearrangements, duplications, deletions, inversions, and translocations (**Figure 1**). SVs impacted *CDKN2A* and *CDKN2B* in 14 samples; *ERBB2* in 6 samples; *SMAD4* in 3 samples; *MDM2, ARID1A,* and *CCNE1* in 2 samples each; and *TP53* in 1 sample. Furthermore, there were 3 tumor samples that demonstrated SVs involving *TP53BP1*, the *TP53* binding protein 1. Additional recurrent SVs affected oncogenes such as *LRP1B* (12 samples), *PTPRD* (12 samples), and *ERBB4* (6 samples). SVs involving *CDK12*, an emerging therapy target (30), were also identified in 8 tumor samples (15%) (**Figure 1**). These included 3 samples with *CDK12* fusions partnered with *CWC25, RAB5B,* and *STAT3* and 5 samples with other *CDK12* SVs including two cases with inversions, two others with a duplication, and one case with a complex rearrangement. SVs impacting genes of the homologous recombination deficiency (HRD) pathway were also observed and included fusion events in *ATM, FANCL,* and *ATR,* an inversion in *PALB2*, a deletion in *ATM*, and a duplication in *RAD51B*.

Targetable alterations were categorized based on 2 criteria: (1) the highest level of evidence for targetability by available treatment agents, and (2) the type of variants, which included oncogenic mutations, amplifications, fusions, TMB-H, and MSI-H. Sixty-two percent (n=32/52) of the samples in the cohort harbored at least one targetable genomic variant (**Figure 2A** and **Figure 2B**). Of those 32 samples, 23 harbored 2 or more targetable alterations A total of 88 targetable variants were identified, 15 (17%) of which had level 1 (L1) treatment correlation evidence (**Figure 2C**). This included *ERBB2* amplification in 5 samples, TMB-H in 4 samples, MSI-H in 2 samples, and targetable fusions (*ISG20L2::NTRK1* and *ADGRV1::NTRK3)* in liver metastases from 2 different patients. Other targets included fusions in *NRG1, ALK, MET* and *CDK12* (as mentioned above). Of the samples with multiple targets (n=23), two samples had concurrent MSI-H and TMB-H, and one TMB-H sample had a concurrent *ADGRV1::NTRK3* fusion as mentioned previously.

**Figure 2.**
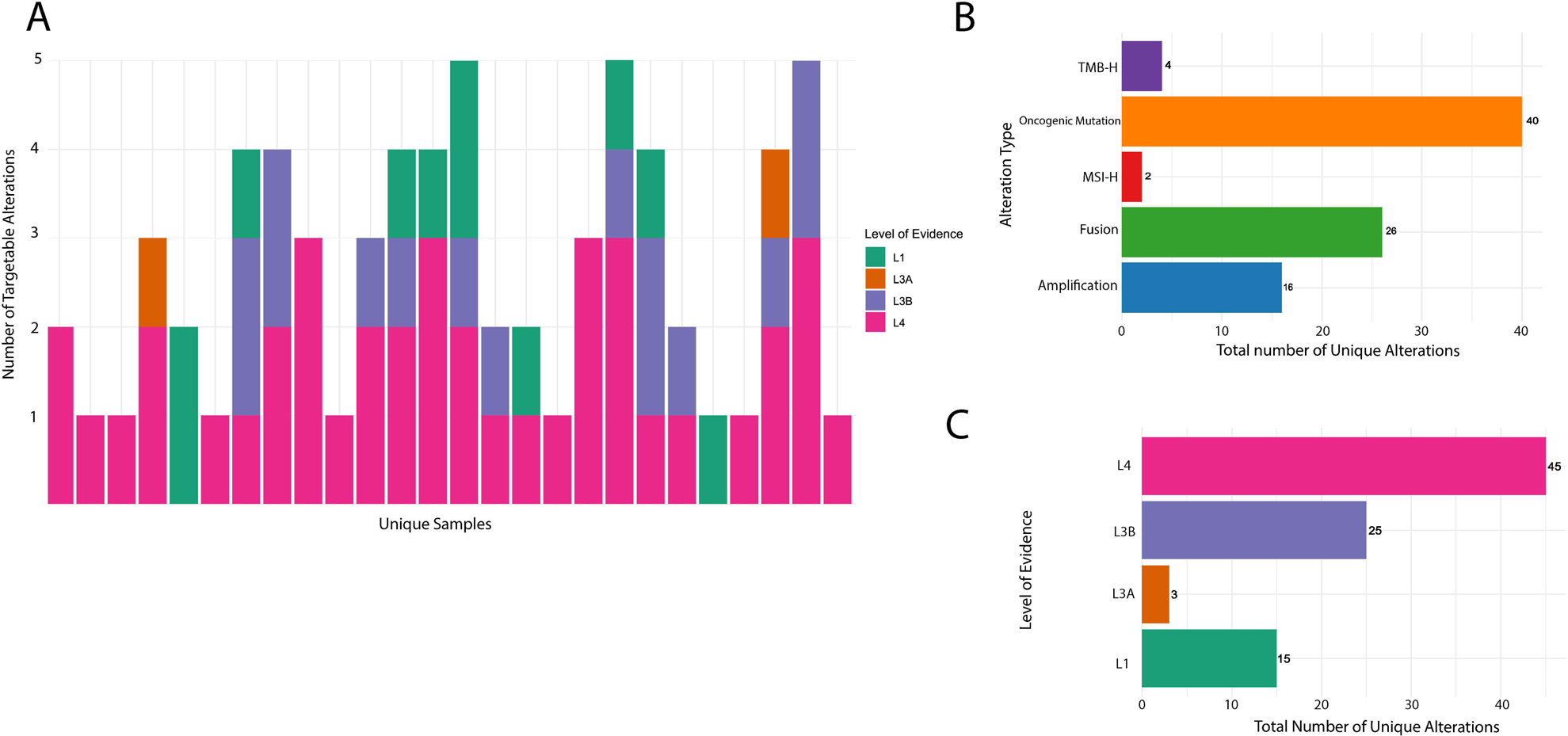
Integrated whole genome and transcriptome analysis of esophagogastric adenocarcinoma reveals significant therapeutic targets. Number of targets and level of evidence for targetability in each sample (A), types of targetable variants in the study cohort (B), and level of evidence for targetability (C) are highlighted. Abbreviations: MSI-H = Microsatellite instability high; TMB = Tumor mutational burden; L1 = Level 1 evidence; L3A = Level 3A evidence; L3B = Level 3B evidence; L4 = Level 4 evidence.

### Identification of a Subset of EGAC with Hypermutator and Tandem-Duplicator Phenotypes

Four samples exhibited TMB-H based on the measurement of whole-genome TMB, two of which also exhibited MSI-H. The first sample featured an *HNF1A* cancer hotspot mutation(31) and SBS molecular signatures related to aging (SBS1 and SBS5). The second sample displayed an *MLH1* frameshift variant (p.T116fs*20) and a high prevalence of SBS signatures associated with mismatch repair (MMR) (*i.e.,* SBS26 and SBS34), accounting for 68% of the mutational burden. The third and fourth samples consisted of two metastatic liver samples collected from the same patient one year apart. One sample had a whole genome TMB of 12.2 mut/mb and a coding (exomic) TMB of 6.7 mut/mb and the other one harbored a whole genome TMB of 21.2 mut/mb and a coding TMB of 16.3 mut/mb (acquired mutator phenotype)(32) (**Figure 3A**). The latter sample also exhibited SBS signatures associated with damage by reactive oxygen species (ROS) and 5-fluorouracil (5-FU) treatment (SBS17b and SBS18), contributing to 31% of the TMB. This correlates with the documented treatment with FOLFOX (5-FU, oxaliplatin, folinic acid) received in the interim. Histologic evidence of treatment effect can be seen with nuclear irregularity, hyperchromasia, and cytologic atypia in both samples.

**Figure 3.**
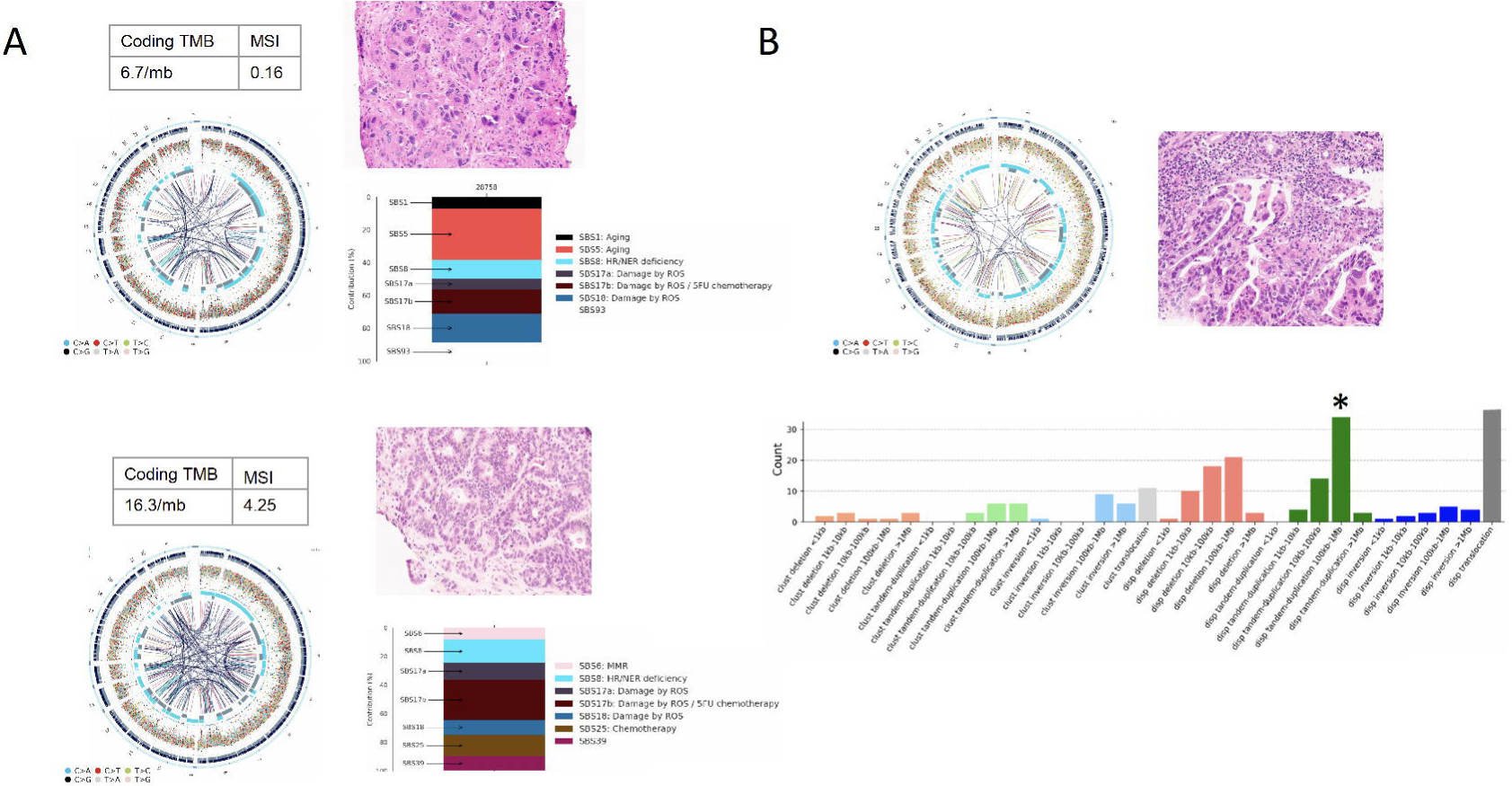
Acquired hypermutator phenotype and *CDK12*-type genomic instability identified in esophagogastric adenocarcinoma. (A) One case demonstrated an acquired hypermutator phenotype. The treatment naïve primary tumor (top) has a coding TMB of 6.7 mut/mb and higher contribution of molecular signatures SBS5 associated with aging. In contrast, the metastasis (bottom) demonstrates a high TMB, 16.3 mut/mb and a predominant contribution of signatures SBS17b associated with 5-fluorouracil chemotherapy. Both tumor samples are microsatellite stable. Corresponding circos plots and histology images (H&E stain) are shown. (B) An example of esophagogastric adenocarcinoma with *CDK12*-type genomic instability associated with a *CDK12* structural variant. Genome circos plot and histology image (H&E stain) are shown. A total of 216 SVs were present with enrichment of large tendem duplications as illustrated in the SV signatures bar plot. Abbreviations: SBS = Single base substitution; MSI = Microsatellite instability high; SV = Structural variant, TMB = Tumor mutational burden, mb = megabase, H&E = Hematoxylin and eosin.

We identified two samples with *CDK12*-type genomic instability (33–36), characterized by an excess of large tandem duplications (100Mb-1Mb) and associated *CDK12* fusions with *STAT3* and *RAB5B* in 2 separate cases (as mentioned earlier). The genomic characteristics of the case with a *CDK12::RAB5B* fusion are illustrated in **Figure 3B**. Further, we employed a whole genome-based classifier of HRD (37, 38) and identified one case with a high confidence HRD score (>0.50) supported by increased small deletions (1Kb-10Kb) in rearrangement signatures. The tumor sample harbored a *BRCA2* p.K503fs*6 somatic biallelic frameshift variant, most likely resulting in the HRD phenotype. Three other samples had an indeterminate HRD score (0.10-0.50) characterized by a *KMT2D* frameshift variant, mutations in the HRD pathway including a deleterious somatic *FANCA* frameshift variant, and structural variants in *ATR, CHEK2, FANCE*, and *FANCM*, and biallelic *RAD51B* deleterious complex rearrangement.

## Discussion

While the incidence of EGAC is decreasing in the United States, these malignancies remain deadly with a poor prognosis. Developments in treatments have only marginally improved survival, underscoring the need for a deeper understanding of the genomic underpinnings and molecular targets of these malignancies. We evaluated the potential translational value of integrating state-of-the-art WGS and RNA-seq data analysis with clinico-pathological information and demonstrated relevant biomarkers and therapeutic targets in EGAC including SVs impacting homologous recombination repair genes, novel gene fusions with *NTRK*, and TMB-H tumors with an acquired mutator phenotype (**Figure 4**). Overall, 62% of the tested samples harbored targetable mutations or molecular phenotypes, 23% of which had Level 1 evidence. These variants included TMB-H and MSI-H, which are treatable with immune checkpoint inhibitors (ICIs). In patients with gastric cancer, recent studies have demonstrated an improved response to first-line combination therapy with pembrolizumab and chemotherapy particularly when both TMB and MSI are high (39). Similarly, a better overall response rate to immunotherapy and improved overall survival has been observed in tumors harboring ≥14.3 mut/mb (40). Our study highlights a subset of EGAC cases with distinct molecular phenotypes that may respond favorably to immunotherapy. Interestingly, one case exhibited a more than two-fold increase in coding TMB following recurrence after chemotherapy. The cause of this temporal heterogeneity in TMB has not yet been elucidated in the literature. However, the possibility of spatial TMB heterogeneity was ruled out in a study by Zhou *et al*. (32).

**Figure 4.**
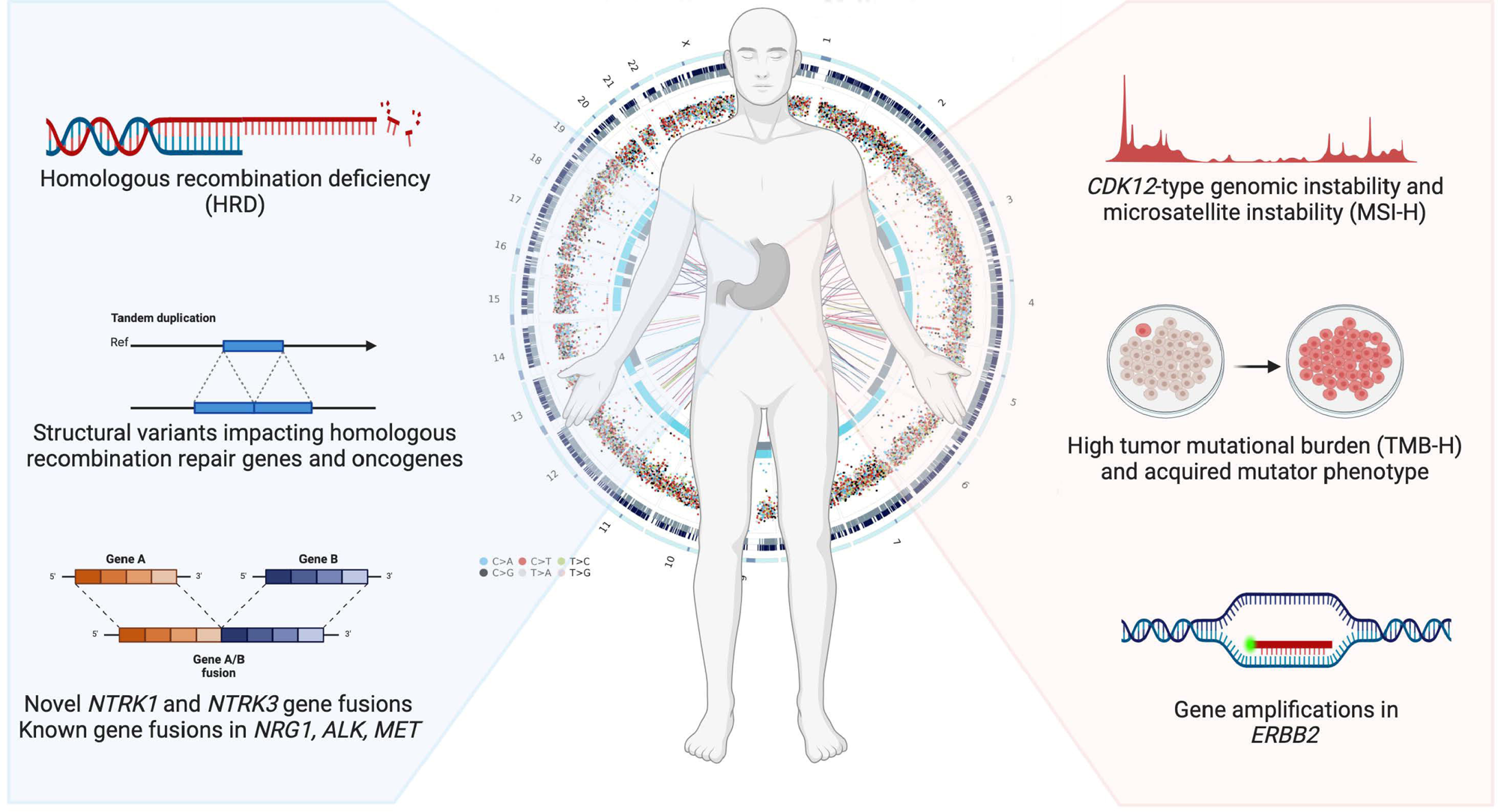
Unearthing relevant biomarkers and therapeutic targets in esophagogastric adenocarcinoma. Highlights of integrated whole-genome and transcriptome sequencing analysis of 52 tumor samples from 47 patients. Overall, 62% of samples harbored targetable mutations or clinically relevant molecular phenotypes.

The identification of novel targetable fusions in EGAC was another significant finding. We identified gene fusions in *NTRK1* and *NTRK3*, which are targetable by tropomyosin receptor kinase (TRK) inhibitors (41). Although fusions involving these genes have been found in multiple cancer types, including colorectal, lung, gastric, and rare cancers(42, 43), we are first to describe the fusion partners in EGAC as verified using the Mitelman Database of Chromosome Aberrations and Gene Fusions in Cancer (44). Other targetable fusions involving *MET* (45) and *NRG1*(46) were also identified.

*CDK12*-type genomic instability has been observed in other cancers, especially prostate, and has demonstrated favorable responses to PD-1 inhibition and poly ADP ribose polymerase (PARP) inhibitors(47–49). This highlights the potential utility of these drugs in treating a subset of EGAC with *CDK12* instability. Three samples with novel fusions involving *CDK12* were identified within our cohort, and two of them demonstrated genomic instability secondary to a large tandem duplication phenotype as described by Sokol *et al*. (33). In contrast, the HRD phenotype, also targetable with PARP inhibitors and sensitive to platinum-based therapy, was only seen in one case. This finding is consistent with the reported low frequency of HRD in esophagogastric cancer (50).

Complex structural variant patterns - undetectable by other molecular assays, were found in *ATM*, Fanconi anemia genes (*FANCA, FANCB, FANCL*), and *RAD51B*. Despite the low HRD score in these tumors, follow-up of cases with these aberrations may be worth pursuing in the event there is treatment response to platinum-based therapy or PARP inhibitors (51, 52).

We also demonstrated additional value of WGTS by uncovering structural variants (SVs) impacting cancer genes, which may help identify potential drivers and further our understanding of these tumors. For example, most alterations in *LRP1B* that we detected were SVs. This gene is known to have an oncogenic role in gastric cancer and is mutated in 27% of cases on targeted panel sequencing datasets (53). Other examples of genes impacted by SVs in our study include *FHIT, ERBB4, PTPRD*, and *CDK12*. All of these have been reported to play roles in cancer, functioning either as oncogenes (*ERBB4* and *CDK12*) or tumor suppressor genes (*FHIT* and *PTPRD*)(54–58). Of additional value, *CDK12* is emerging as a potentially targetable marker in prostate, breast, and ovarian cancers, among others (34, 59, 60).

Previous WGS studies have been published on esophageal cancers, mostly studying esophageal squamous cell carcinoma and esophageal adenocarcinoma in the background of Barrett mucosa^58^. Very few studies have been reported on gastric adenocarcinoma and combined cohorts of gastric and esophageal carcinomas^7,59,60^. In this context, our retrospective study offers valuable insights into the molecular underpinnings and detection of additional targets in EGAC (**Figure 4**) by employing novel WGTS analytic tools.

## Conclusions

Esophagogastric adenocarcinoma remains an aggressive disease with poor patient outcomes. In an attempt to identify novel targets and clinically relevant molecular phenotypes, we characterized the entire genome and transcriptome of this disease. Targetable oncogenic events were detected in 62% of tumors, emerging molecular phenotypes with potential clinical relevance were identified, and unique structural variants in relevant cancer genes were discovered. Our findings support the exploration of integrative whole genome and transcriptome sequencing to expand the potential therapeutic targets in esophagogastric adenocarcinoma, a malignancy with dismal prognosis.

## Data Availability

All data produced in the present study are available upon reasonable request to the authors

## List of Abbreviations

GEJ: Gastroesophageal junction
SEER: Surveillance, epidemiology, end results
EGAC: esophagogastric adenocarcinomas
MSI: microsatellite instability
MSI-H: high microsatellite instability
WGTS: whole genome and transcriptome sequencing
WCM: Weill Cornell Medicine
IRB: institutional review board
EMR: electronic medical record
WGS: whole genome sequencing
SNVs: single nucleotide variants
Indels: insertions/deletions
SVs: structural variants
SBS: single base substitution
TMB: tumor mutational burden
HRD: homologous recombination deficiency
TMB-H: high tumor mutational burden
RNA-seq: -RNA sequencing
FFPE: formalin-fixed paraffin-embedded
CNVs: copy number variants
ROS: reactive oxygen species
5-FU: 5-fluorouracil
TRK: tropomyosin receptor kinase

## Ethics approval and consent to participate

All patients in this study signed a written informed consent as part of the IRB–approved protocol No. 1305013903 at Weill Cornell Medicine.

## Consent for publication

This manuscript does not contain any individual person’s data in any form.

## Availability of data and materials

The data presented in this study are not publicly available due to data size and patient privacy. However, the datasets used/or analysed during the current study are available from the corresponding author upon reasonable request.

## Authors’ contributions

Concept and design of the study: Annika Windon, Majd Al Assaad and Juan Miguel Mosquera

Patient enrollment: Manish Shah and Allyson J. Ocean

Collection of clinical data and metadata extraction: Jyothi Manohar, Marvel Tranquille, Annika Windon and Majd Al Assaad

Histopathology review: Annika Windon, José Jessurun, Nicole Mendelson, Erika Hissong and Kenrry Chiu

Lab experiments: Michael Sigouros, Justin Mclee and Marvel Tranquille

Computational analyses and sequencing data management: Kevin Hadi, Aditya Deshpande, Minal Patel, Juan S. Medina-Martínez, Andrea Sboner and Olivier Elemento

Writing of first draft of the manuscript: Annika Windon and Majd Al Assaad

Writing and editing of the revised manuscript and study supervision: Juan Miguel Mosquera

All authors reviewed data for publication and approved the manuscript.

## Acknowledgements

Project support for this research was provided in part by the Center for Translational Pathology (Ruben Diaz, Leticia Dizon, Bing He) from the Department of Pathology and Laboratory Medicine at Weill Cornell Medicine. The authors thank Ahmed G Elsaeed for project support with data extraction. Figure 4 was created with BioRender.com.

